# Post-COVID-19 syndrome: retinal microcirculation as a potential marker for chronic fatigue

**DOI:** 10.1101/2022.09.23.22280264

**Authors:** Sarah Schlick, Marianna Lucio, Alexander Bartsch, Adam Skornia, Jakob Hoffmanns, Charlotte Szewczykowski, Thora Schröder, Franziska Raith, Lennart Rogge, Felix Heltmann, Michael Moritz, Lorenz Beitlich, Julia Schottenhamml, Martin Herrmann, Thomas Harrer, Marion Ganslmayer, Friedrich E. Kruse, Robert Lämmer, Christian Mardin, Bettina Hohberger

## Abstract

Post-COVID-19 syndrome (PCS) summarizes persisting sequelae after infection with the severe-acute-respiratory-syndrome-Coronavirus-2 (SARS-CoV-2). PCS can affect patients of all covid-19 disease severities. As previous studies revealed impaired blood flow as a provoking factor for triggering PCS, it was the aim of the present study to investigate a potential association of self-reported chronic fatigue and retinal microcirculation in patients with PCS, potentially indicating an objective biomarker.

A prospective study was performed, including 201 subjects: 173 patients with PCS and 28 controls. Retinal microcirculation was visualized by OCT-Angiography (OCT-A) and quantified by the Erlangen-Angio-Tool as macula and peripapillary vessel density (VD). Chronic Fatigue (CF) was assessed with the variables ‘Bell score’, age and gender. The VD in the superficial vascular plexus (SVP), intermediate capillary plexus (ICP) and deep capillary plexus (DCP) were analyzed considering the repetitions (12 times). Taking in account of such repetitions a mixed model was performed to detect possible differences in the least square means between different groups of analysis.

An age effect on VD was observed between patients and controls (p<0.0001). Gender analysis yielded that women with PCS showed lower VD levels in SVP compared to male patients (p=0.0015). The PCS patients showed significantly lower VD of ICP as compared to the controls (p=0.0001, [CI: 0.32; 1]). Moreover, considering PCS patients, the mixed model reveals a significant difference between chronic fatigue (CF) and without CF in VD of SVP (p=0.0033, [CI: -4.5; -0.92]). The model included age, gender and the variable ‘Bell score’, representing a subjective marker for CF. Consequently, the retinal microcirculation might be an objective biomarker in subjective-reported chronic fatigue of patients with PCS.

## 1. Introduction

The severe-acute-respiratory-syndrome-Coronavirus-2 (SARS-CoV-2) first observed in Wuhan, Hubei Province, China, 2019 [1] has been declared as public health emergency of international concern in January 2020 from the world health organization (WHO). [2] In March 2020 it reached levels of a worldwide pandemic with impact on social life, economy and health care system. [3] The acute Coronavirus disease 2019 (COVID-19) caused a number of pneumonia cases [1] and can lead to complications like respiratory and multiorgan failure. [3] By February 18, 2022, the virus spread to over 418 million cases with over 5.8 million deaths (numbers from WHO). [4, 5] Apart from the acute COVID-19 disease, the post-COVID syndrome (PCS) can arise afterwards. PCS is defined as persisting symptoms for more than 12 weeks after infection with the virus (S1 guideline; AWMF online). Continuing symptoms for more than 4 weeks after infection are defined as Long-COVID or post-acute sequelae of COVID-19. [6]

The most common symptoms reported in studies are chronic fatigue (CF) and dyspnea (i.e., shortness of breath). Persistent symptoms could be neurocognitive impairments (brain fog, loss of attention), autonomic symptoms (chest pain, palpitations, tachycardia), gastrointestinal issues, musculoskeletal problems (myalgia), smell and taste dysfunction, cough, headache, and hair loss. [5-7] Studies reported that PCS affects even people with moderate acute COVID-19 who didn’t require hospital care during the acute stage. [5, 8, 9] The absolute number of patients with PCS goes along with the shape and amplitude of the pandemic curve showing the risk of PCS for individual health, healthcare system and economy. [10] Studies revealed that more than half of the patients with COVID-19 infection reported PCS-symptoms. [11] Others postulate that 15% of all COVID patients [6] and 50-70% of hospitalized patients suffer from PCS. [7]

Pathogenesis of PCS is still elusive. Recent studies revealed viral persistence, enduring texture damage including endotheliopathy, impaired microvasculature, hypercoagulation, thrombosis, neutrophil extracellular traps (NETs), chronic immune dysregulation, dysregulation of the renin-angiotensin-aldosterone-system (RAAS) and hyperinflammation/autoimmunity as possible pathomechanisms of PCS. [6, 12-14] It is assumed that autoimmunphenomena [15] including the generation of functional active autoantibodies [5, 16] are involved in the pathogenesis of PCS with potential different PCS subgroups. [5, 6] Studies examined increased D-dimer levels up to 4-month post-acute infection in approximately 25% of patients. The mechanisms of these persistent procoagulant effects in PCS are not clarified at this point of time. [17] Endotheliopathy and elevated plasma markers of endothelial cell activation have been recognized in patients with severe COVID-19. Studies investigated a persistent endotheliopathy in patients with PCS and it is associated with enhanced thrombin generation potential independently of ongoing acute phase response or NETosis. [17] Autopsy studies revealed that alveolar capillary microthrombi were 9-times more prevalent in patients with COVID-19 compared to patients with influenza. [18] Fatigue is one of the most common symptoms of PCS which emphasizes its impact on individual health, health care system and economics. [6, 11, 19] The study “Assessment and characterization of post-COVID-19 manifestations” revealed that fatigue was reported with 72.8% as the most common symptom. [20] Sudre et al related to data from the COVID symptom study app and concluded that “self-reported fatigue is the commonest complaint in a large group of Long-COVID patients”. [21, 22] Apart from PCS, fatigue is a possible postinfectious consequence after infection with infectious diseases caused by e.g., Epstein-Barr virus (EBV), humanherpesvirus 6, influenza virus or Coxiella burnetii. However, a much higher rate of post-COVID fatigue was observed compared to e.g. post-EBV, Q-fever, considering a similar interval. [11, 23] Fatigue in PCS shows phenotypical similarity to chronic fatigue syndrome (CFS), also called myalgic encephalomyelitis (ME), which is often induced by an infectious agent. [6, 19] ME/CFS is defined as persistent fatigue for 6 month or longer. Viral and bacterial infections are thought to be the cause of ME/CFS, but pathophysiology is still elusive. [11] An association between fatigue in PCS and laboratory markers of inflammation and cell turnover (leukocyte, neutrophil or lymphocyte counts, neutrophil-to-lymphocyte ratio, lactate dehydrogenase, C-reactive protein) or pro-inflammatory molecules (IL-6 or sCD25) were not observed until now. [11] Thus, it would be of interest to establish an objective marker for the patients’ self-reported fatigue.

The eye as window to the human body can be used as “diagnostic window” for several systemic disorders. Diabetes [24, 25], arterial hypertension [26, 27] and several metabolic diseases [25, 28] show their first sign within the retina. The retinal capillary system represents the microcirculation in the whole human body; thus, retinal capillary disorders might represent the whole human microcirculation. Retinal macular and peripapillary capillary plexi can be visualized by Optical-Coherence-Tomography-Angiography (OCT-A) and quantified by the Erlangen-Angio-Tool (EA-Tool). [5, 29-31] OCT-A is easy to handle and measures non-invasively without any contact to the human eye. It measures differences in the speckle pattern of backscattered light in two or more repeated scans. These differences are caused by moving particles like red blood cells (RBC). [32] The aim of this study was to investigate the association of self-reported chronic fatigue and retinal microcirculation in patients with PCS, potentially indicating an objective biomarker.

## Material and Methods

### Participants

One-hundred seventy-three patients with post-COVID (age: 39.7±12, gender: 109 female; 64 male) and 28 controls (age: 29.2±12, gender: 20 female; 8 male) were recruited at the Department of Ophthalmology, University of Erlangen-Nürnberg, Friedrich-Alexander-Universität Erlangen-Nürnberg (FAU). Post-COVID syndrome was defined as persisting symptoms for more than 12 weeks after infection with the virus according to the S1-Guidline. [6] SARS-CoV2 infection was confirmed by a positive real-time, reverse-transcription-polymerase-chain-reaction-test. Time after COVID-19 was 231±111 days. Post-COVID symptoms were CF (92%), impaired concentration (83%), hair loss (63%), POTS (19%), and subjectively colder hands (12%). All eyes showed no local or systemic disorders with retinal affection. The patients underwent an ophthalmic examination including measurement of best-cured visual acuity (BCVA), non-contact intraocular pressure (IOP), measurement of axial length (IOL Master, Zeiss, Oberkochen, Germany) and OCT-angiography (see below in detail). Anamnestic data including self-reported chronic fatigue were recorded. In addition, a subgroup of patients with post-COVID assessed their self-reported fatigue in a chronic fatigue score (Bell Score). All patients signed a written informed consent form. The study was approved by the local ethics committee and performed in accordance with the tenets of the Declaration of Helsinki.

### OCT-A

OCT-A (Heidelberg Spectralis II, Heidelberg, Germany) is a diagnostic technique visualizing the retinal microcirculation of the macula and peripapillary region. Retinal macula microvasculature can be subdivided in three layers: superficial vascular plexus (SVP), intermediate capillary plexus (ICP) and deep capillary plexus (DCP). All OCT-A scans have an angle of 15° covering a size of 2.9 mm x 2.9 mm with a lateral resolution of 5.7 µm/pixel.

The OCT-A data were exported by the SP-X1902 software (prototype software, Heidelberg Engineering, Heidelberg, Germany) and analyzed by the Erlangen-Angio-Tool (EA-Tool) software, which is coded in MATLAB (The MathWorks, Inc., Natick, USA, R2017b). Studies revealed a high reliability and reproducibility of EA-Tool. [33] For the analysis, the VD was computed in 12 sectors for macular. Moreover, the overall VD was computed as a mean over the sectors. In addition, the Anatomic Positioning System (APS, part of Glaucoma Module Premium Edition (GMPE), Heidelberg Engineering, Heidelberg, Germany) was implemented into the EA-Tool. This feature aligns all OCT-A scans according to their individual Fovea-to-Bruch’s Membrane Opening-Center axis (FoBMOC) to allow for a better comparison of different scans. This FoBMOC axis is defined by the fovea and the center of the Bruch’s Membrane Opening. [5]

### Statistical Analysis

The data were analyzed by different mixed models (SAS version 9.4, Institute Inc., Cary, NC, USA) taking into consideration the repetitions of the eyes (12 times) for each sector of macula OCT-A scans. In the first model we compared the PCS versus controls patients, the variable was set as independent variable. In the second one we excluded the control group. The independent variable was chronic fatigue. We estimated the least squares means (LS means) that corresponded to the specified effects for the linear predictor part of the model and the relative confidence limits. LS means are closer to reality and represent even more real data, when cofactors occur, compared to means. Age and gender were introduced as covariates in both models. In the second model we also added the bell score as predicative variable. The p-values (the α value is set at 0.05) are presented with the respective confidence interval limit (CL). All the CL and the p-values for the multiple comparisons are adjusted with Tukey-Kramer.

## Results

LS means of overall VD were 29.97±0.06 (SVP), 21.96±0.05 (ICP), and 23.62±0.06 (DCP) in patients with PCS. Controls showed LS means of overall VD of 30.13±0.19 (SVP), 22.62±0.17 (ICP), and 23.73±0.19 (DCP).

A significant effect of age and gender were observed on VD of SVP, ICP, and DCP, respectively (p<0.0001). Considering an influence of age, we observed that with increasing age a decreasing VD of SVP, ICP, and DCP was observed (figure 1). Estimate values were -0.06 (SVP), -0.06 (ICP), and -0.07 (DCP) in patients with PCS.

**Figure 1:**
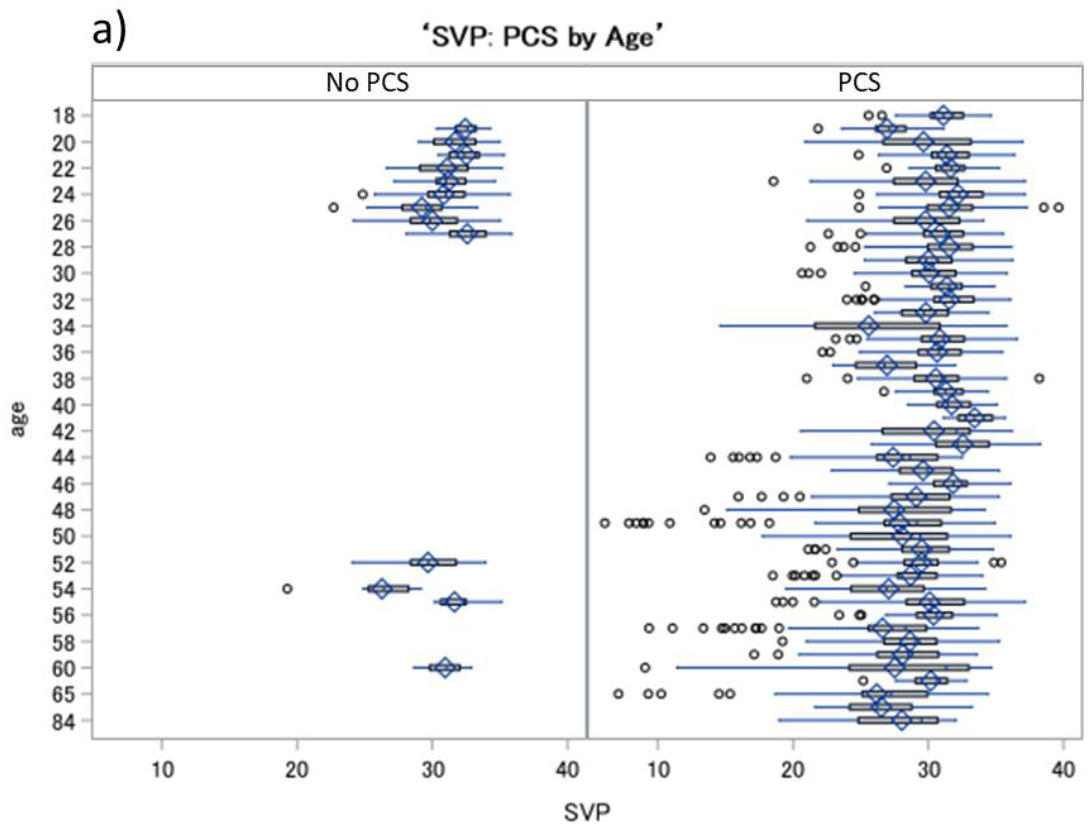
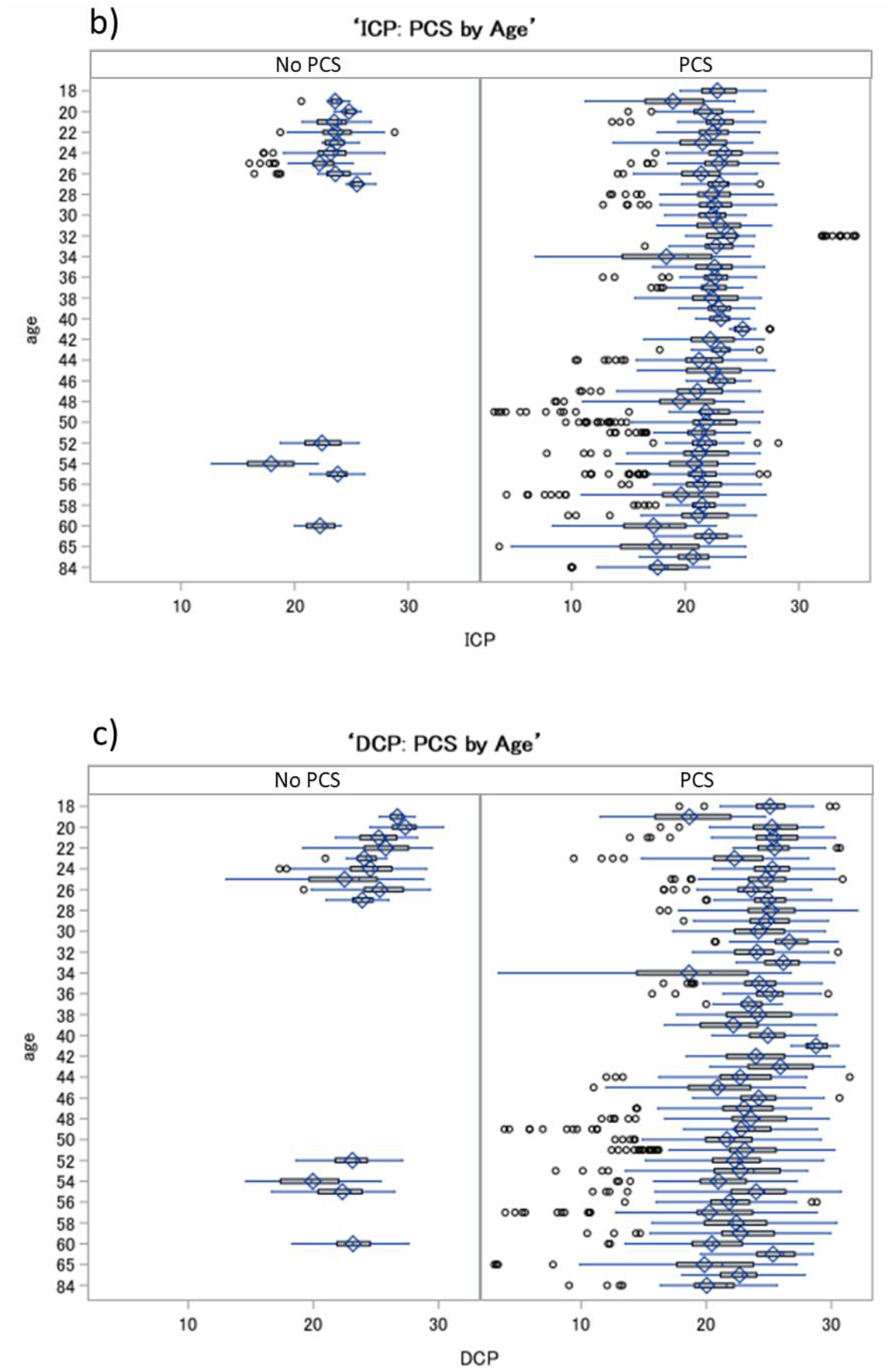
Boxplots of VD in SVP (a), ICP (b), and DCP (c) for patients with post-COVID syndrome (PCS) and controls considering the cofactor age: a decrease of VD in all retinal layers was observed with increasing age in PCS and controls; SVP-superficial vascular plexus, ICP – intermediate capillary plexus, DCP – deep capillary plexus.

The correlation of pairs of each VD of SVP, ICP, and DCP is plotted considering gender in patients with PCS in Figure 2. Female showed lower levels for each comparison pairs of retinal layers. Especially, a significant decreased (Type 3 Tests of Fixed Effects) VD of SVP of female patients with PCS was observed compared to male (LS-Mean difference = 1.05, [CI: 0.41; 1.69, p=0.0015; Figure 3) with increase of age.

**Figure 2:**
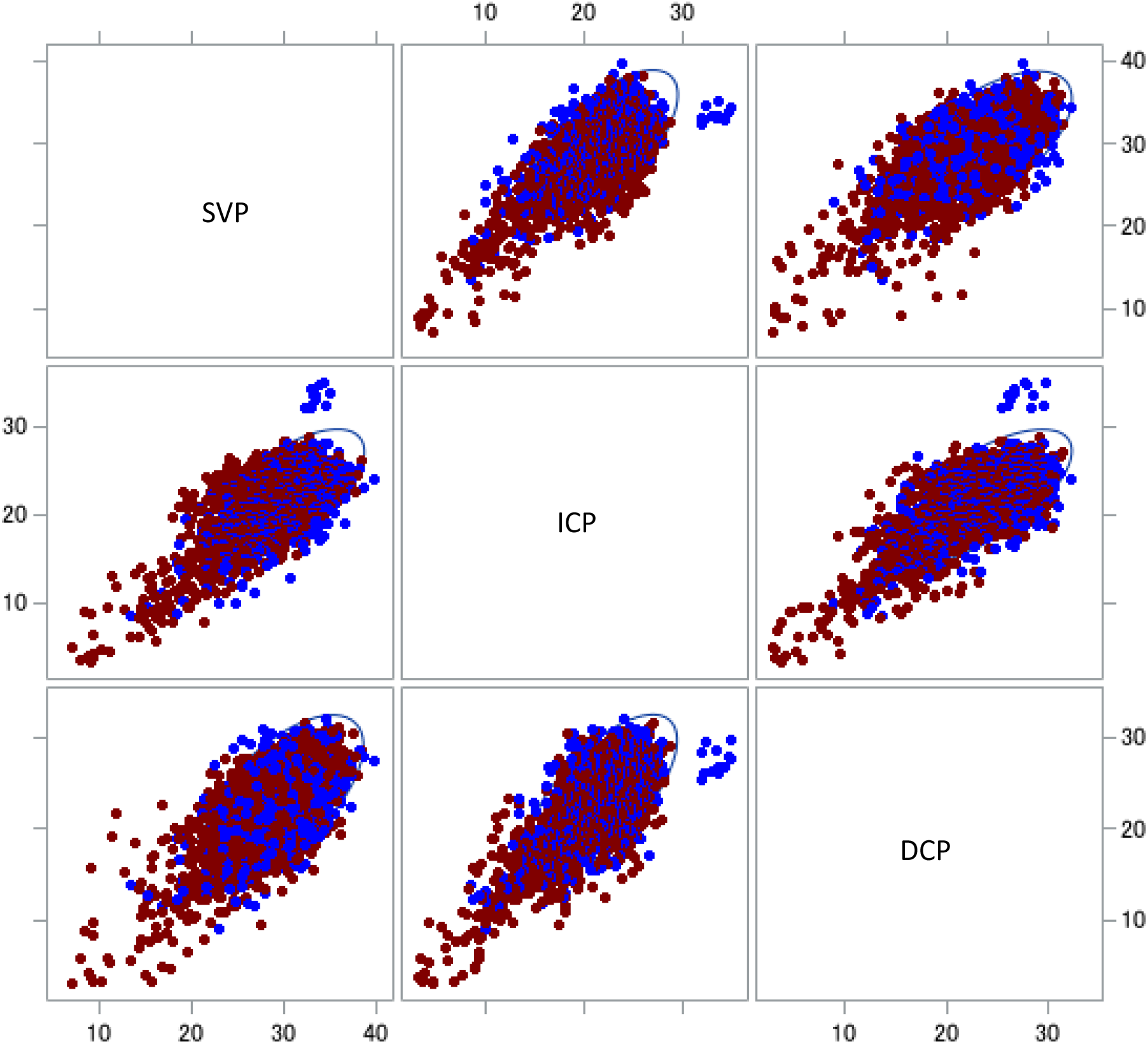
Scatter plot of Vessel density differences in patients with post-COVID syndrome (PCS) regarding gender: matrix showing the relation between each vessel density of SVP, ICP, and DCP of patients with PCS, respectively. Each pair of vessel density is colored by gender (male, blue; female, red), with 95 % prediction ellipse; Female patients with PCS and CF showed lower VD data; SVP – superficial vascular layer; ICP – intermediate capillary plexus; DCP – deep capillary plexus.

**Figure 3:**
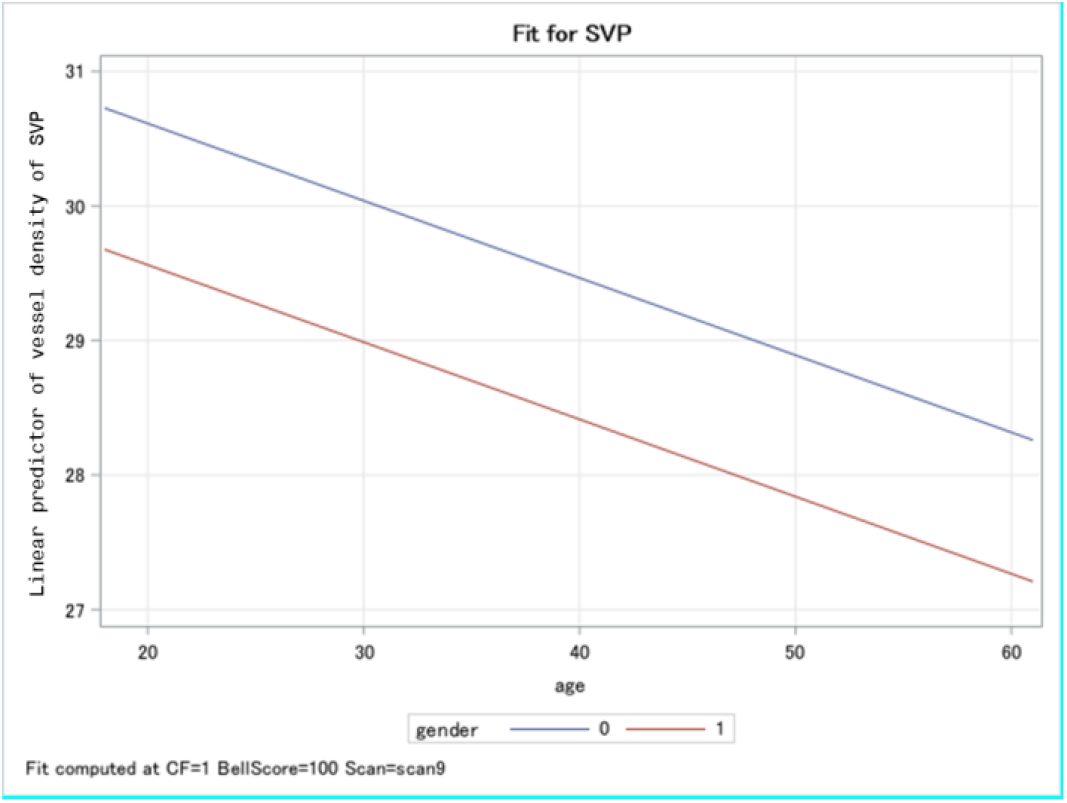
Panel plot that overlays the predicted values for males (blue line) and females (red line) for the Vessel Density of superficial vascular layer (SVP) in patients with post-COVID symptoms: Women patients showed significant lower VD in SVP levels compared to men with increasing of the age.

The analysis using a mixed model with 12 repetitions, which was corrected for age and gender, yielded a significant impaired VD in ICP (p=0.0001), yet not in SVP and DCP (p>0.05) in patients with PCS compared to controls (Table 1).

**Table 1:** Differences of least square means of vessel density in SVP, ICP and DCP in patients with Post-COVID (PCS, 1) compared to controls. SVP - superficial vascular plexus, ICP – intermediate capillary plexus, DCP – deep capillary plexus.

| Differences of Least Squares Means of Vessel Density of patients with PCS compared to controls |  |  |  |  |  |  |  |  |  |
| --- | --- | --- | --- | --- | --- | --- | --- | --- | --- |
|  | Effect | Controls | _PCS | Estimate | SE | t Value | Pr > \|t\| | Lower | Upper |
| **SVP** | Post-COVID | 0 | 1 | 0.16 | 0.2 | 0.81 | 0.42 | -0.23 | 0.55 |
| **ICP** | Post-COVID | 0 | 1 | 0.66 | 0.2 | 3.84 | 0.0001 | 0.32 | 0.1 |
| **DCP** | Post-COVID | 0 | 1 | 0.11 | 0.2 | 0.54 | 0.59 | -0.2855 | 0.5 |

Instead, in the PCS patients the complete model (including age, gender and bell score variables) reveals a significant difference between chronic fatigue (CF) and without CF in VD of SVP (p=0.0033, [CI: -4.5; -0.92]), table 2.. Any notable differences were not observed for the other retinal layers (p>0.05). Patients with PCS and CF showed LS means of VD of 30.3±0.28 (SVP), 21.89±0.25 (ICP), and 23.08±0.24(DCP). Patients with PCS without CF showed LS means of VD of 27.59±0.91 (SVP), 21.58±0.84 (ICP), and 23.69±0.8 (DCP).

**Table 2:** Differences of Least Squares Means of VD of SVP, ICP, and DCP in patients with Post-COVID Syndrome (PCS) with chronic fatigue (CF, 1) and without CF (0): A notable significant effect was observed in SVP (p=0.0033).

| Differences of Least Squares Means of Vessel Density of PCS patients with CF (1) and without CF (0) |  |  |  |  |  |  |  |  |  |
| --- | --- | --- | --- | --- | --- | --- | --- | --- | --- |
|  | Effect | CF | _CF | Estimate | SE | t Value | Pr > \|t\| | Lower | Upper |
| SVP | Chronic Fatigue | 0 | 1 | -2.71 | 0.91 | -2.98 | 0.0033 | -4.51 | -0.92 |
| ICP | Chronic Fatigue | 0 | 1 | -0.31 | 0.84 | -0.37 | 0.71 | -1.97 | 1.35 |
| DCP | Chronic Fatigue | 0 | 1 | 0.61 | 0.8 | 0.76 | 0.45 | -0.97 | 2.19 |

## Discussion

Post-COVID-19 Syndrome (PCS) is a challenge for individual health, the health care system, and economy according to its high prevalence in patients worldwide. Leading clinical symptom of PCS is self-reported fatigue as it is one of the most common symptoms in PCS, considering large groups of patients. [6, 11, 19-22] The prevalence of CF is not associated with COVID-19 severity which implicates its potentially high number of patients. [11] As each clinician prefers objective biomarkers in addition to self-reported clinical symptoms, the aim of this study was to investigate the association of self-reported chronic fatigue and retinal microcirculation in patients with PCS, potentially indicating an objective biomarker. An age effect on VD was observed in patients and controls (p<0.0001). Gender analysis yielded that especially women with PCS showed lower VD levels in SVP compared to male patients (p=0.0015). Previous studies revield that women show a ratio of PCS of 3:1 ratio compared to male. [34, 35] In addition, the present data go conform with clinical observations that women with PCS had a higher probability of fatigue and anxiety/depression throughout 6-month-follow up. [36, 37] Patients with PCS showed significant lower VD of ICP compared to controls (p=0.0001, [CI: 0.32; 1]), considering the age and gender effect. Instead, in the PCS patients the mixed model reveals a significant difference between chronic fatigue (CF) and without CF in VD of SVP (p=0.0033, [CI: -4.5; -0.92]). In the model was included age, gender, and the variable ‘Bell score’, representing a subjective marker for CF. The variable ‘Bell score’ was always significant for each VD. Thus, the eye as window in the human body might offer a diagnostic option by measuring retinal microcirculation objectively in self-reported CF of patients with PCS.

Up to date there is no uniform consensus of a definition for CF in PCS: It is assumed that the label CF/post-COVID-fatigue, going in line with definitions of post-infection fatigue, should be applied under following conditions: a dominant symptom, chronic, disabling as it prevents pre-illness activities and duties, intensified after mental and/or physical activity (post-exertional malaise, PEM) [38-40], persistent for 6 month or longer (3 month in children and adolescents), occurring during confirmed acute COVID-19 and without symptom-free interval since onset. [41] The unknown nature of PCS and phenotypical similarity to a postinfectious fatigue syndrome lead some studies to suggest a connection to myalgic encephalomyelitis/ chronic fatigue syndrome (ME/CFS). [42-44] Post exertional malaise (PEM), a leading symptom of ME/CFS, describes worsening symptoms after low or moderate daily activity for several hours or weeks. This burden has also been found in PCS patients. [39, 40] ME/CFS is defined by at least six months of PEM and exhaustion, additionally suffering frequently from orthostatic intolerance or cognitive decline. Patients loose ability to engage in pre-illness levels concerning social life, work, or school duties. [44] Studies revealed that patients react abnormal to stressors like e.g. waking up with an abnormal rise in serum cortisol and heart rate. [45] Women seem to be more affected than men. [46]

CF in PCS exhibits similar incidences in hospitalized and non-hospitalized patients. Fatigue and cognitive impairment assume to endure and may worsen over time in individuals in < 6 months and ≥ 6 months follow-up. [19, 47] If CF in PCS is identified, there should follow an underlying diagnostic. At the moment, CF-diagnostic is based on brief questionnaires to characterize the fatigue state, such as the Calder Fatigue Scale or the SPHERE. These methods try to identify CF in line with the disease-specific recommendations from the National Institute of Neurological Disorders and Stroke Common Data Elements. As CF is often part of a multisymptomatic cluster, SPHERE includes related physical symptoms in the diagnostics and others also include mental health questions (e.g., the Patient Health Questionnaire-9). [41] CF in PCS is associated with marked functional impairment. [19] As a subgroup of patients still exhibited inflammatory markers after the acute COVID-19 infection it has been suggested that hyperinflammation is a cause of CF in PCS. [19] The causal association between specific pro-inflammatory cytokines, mood symptoms and cognitive decline is confirmed. [48, 49] Other post-infectious syndromes (e.g., post-infectious encephalitis) have been associated with inflammatory processes. [50] The pathophysiology of CF remains unresolved. [41]

To the best of our knowledge the present study revealed for the first time an association between CF in PCS by the variable “bell score” and impaired retinal microcirculation in OCT-A, potentially indicating an objective biomarker. OCT-A is able to visualize retinal macular and peripapillary capillary plexi. [5, 29-31] It is easy to handle and measures non-invasively without any contact to the human eye. The technical basis is the recording of a real-time motion signal, based on temporal changes of intravascular moving red blood cells (RBC). [32] If a signal is recorded this retinal pixel is coded by ‘white’ and without any motion it is coded by ‘black’ (binary code). [5] The data can be analyzed with a high reliability and reproducibility with the Erlangen-Angio Tool (EA-Tool). [33] Fine analysis can be done by division of the scan region into 12 sectors (macula) or 4 sectors (peripapillary region) to calculate the overall and sectorial vessel density (VD). The eye as “window” to the human body is representative for several systemic disorders. [24-28] Alveolar capillary occlusion is a characteristic symptom of COVID-19 which can lead in severe cases to respiratory failure as the blood oxygen uptake is limited. [18, 51] Impaired microcirculation can be found in acute COVID-19 as well as in PCS. [5, 52-54] The virus may infect endothelial cells directly via Angiotensin Converting Enzyme 2 (ACE2) which leads to inflammation and fibrosis. [52] The present study investigated female patients with PCS showing lower levels of vessel density (VD) for each comparison pairs of retinal layers (SVP, ICP, DCP) than men. This reinforced impaired microcirculation in female patients with PCS goes in line with results of other PCS studies. [5] The analysis, which was corrected for age and gender, yielded a significant impaired VD in ICP in patients with PCS compared to controls. Interestingly, the Angiotensin Converting Enzyme 2 (ACE2), a serine protease, is located in the retinal layer ICP. There is evidence that SARS-CoV-2 occupies the human body by first binding to the ectoenzyme ACE2, acting as the receptor. Another serine protease is required to prime the viral spike “S” protein for entering the cells. [55] Inclusion of the additional explicative variable ‘Bell score’, representing a subjective marker for CF, in the mixed model, a significant effect was observed in SVP when comparing patients with PCS with CF and without CF. Thus, CF has a significant impact on retinal microcirculation which implicates that there is a potential objective biomarker to illustrate CF and might enable an objective diagnostic option for CF in PCS. The eye as window in the human body might offer a diagnostic option by measuring retinal microcirculation objectively in self-reported chronic fatigue of patients with PCS. It can be assumed that retinal microcirculation can have an impact as diagnostic tool in PCS and additionally might also in related disease like ME/CFS.

## Conclusion

Post-COVID-19 syndrome is a post-infectious disease with multifactorial pathomechanism and symptoms. We could reveal differences in the VD of ICP between control and PCS patients. Considering the PCS patients’ group, we could retrieve differences in VD of SVP between PCS patients with and without CF. As self-reported fatigue is one of the most common symptoms in PCS, the present study yielded that the vessel density of the retinal microcirculation as measured by OCT-A might be an objective biomarker for this subjectively reported symptom.

## Data Availability

All data produced in the present study are available upon reasonable request to the authors

## Author Contributions

Conceptualization, C.M., M.H., M.G., and B.H.; Data curation, C.S., J.H., T.S., F.R., J.S. and B.H. ; Formal analysis, C.S., M.L., L.R., F.H., M.M., L.B. and J.S.; Investigation, C.S., J.H., T.S., F.R., L.R., F.H., M.M., M.G., A.S., A.B., and B.H.; Methodology, C.S., M.L., L.B., M.H., T.H., M.G., R.L., M.Z. and A.G.; Project administration, C.M., J.H., F.K. and B.H.; Software, J.S.; Supervision, C.M. and B.H.; Validation, B.H.; Visualization, M.L., S.S. and B.H.; Writing-original draft, S.S., M.L., and B.H.; Writing-review & editing, C.M.,B.H., M.K., and J.G.; Funding Acquisition, B.H., CM, MG; Resources, C.M., A.G., M.Z., T.H. All authors have read and agreed to the published version of the manuscript.

## Institutional Review Board Statement

The studies involving human participants were reviewed and approved by the ethics committee of the University of Erlangen-Nürnberg (295_20 B). The patients/participants provided their written informed consent to participate in this study.

## Informed Consent Statement

**Informed** consent was obtained from all subjects involved in the study.

## Data Availability Statement

The original contributions presented in the study are included in the article/Supplementary Material.

## Acknowledgments

The present work was performed in fulfillment of the requirements for obtaining the degree Dr. med. for S.S. The Department of Ophthalmology is part of the Universität of Erlangen-Nürnberg, Friedrich-Alexander-Universität Erlangen-Nürnberg (FAU), Germany.

